# Preclinical and Clinical Study of Efficacy of NAOQ19 against SARS-COV2: A Comprehensive Evaluation

**DOI:** 10.64898/2026.01.27.26344593

**Authors:** Saumya Subramanian, Jaishree Garhyan, Vinod Mony, Shrivatsav Pattabiraman, Divya Kanchibhotla

## Abstract

**Background:** More than 6.2 million people have died already from COVID-19. Drug resistance and relapse cases from first generation therapeutics calls for development of new drugs in alternative medicine. Complementary and Alternative Medicines (CAM) that include herbal remedies and phytochemicals are usually not fully integrated into mainstream healthcare systems. The study proposes a CAM remedy, a new polyherbal formulation ‘NAOQ19’ against the SARS-CoV-2.

**Methods:** The present study consists of *invitro* and *invivo* evaluation of NAOQ19 against SARS-CoV-2 infection. First, *invitro* testing of NAOQ19 anti-viral activity was carried out on three relevant cell lines: Vero E6, A549ACE2 and Huh 7.5.1 ACE2TMPRSS2. Next, animal model testing of NAOQ19 was performed in Syrian golden hamsters along with positive control Remdesivir and infection control for 3 days to determine the efficacy and safety of the formulation. Finally, a double blind randomized clinical trial with mild to moderate COVID-19 infected patients were evaluated to test the efficacy of NAOQ19 in human settings.

**Results:** This study demonstrated a strong anti-viral (low EC50) activity in cell culture with live virus and exhibits reduced plaque forming units (high antiviral activity) in the Syrian golden hamster model. Moreover, in the clinical trials, NAOQ19 shows high efficacy demonstrating early recovery and reduced levels of inflammatory biomarkers among COVID-19 infected patients.

**Conclusion:** This novel polyherbal formulation NAOQ19, demonstrates strong anti-viral activity in preclinical and clinical study; thereby proving its candidacy as a low-cost alternative medicine with minimal adverse effects.

## 1. Introduction

Global integration, migration, travel, trade and close interactions between animals and humans have led to emergence of epidemics, endemics and pandemics. Over the years, many pandemics have been caused by zoonotic pathogens that were transmitted to humans [1]. The beginning of the 21^st^ century was marked by two epidemics and one pandemic, all caused by a single family of coronavirus [2]. Coronavirus disease-2019 (COVID-19), caused by SARS-CoV-2, was declared a pandemic by the World Health Organization (WHO) in early 2020 [3]. Since then, the pandemic has spread across 213 countries and territories and infected millions across the globe [4]. As the virus evolved, different variants showed increased infectivity (delta) and increased transmissibility (omicron) [5]. Morbidity rates were highest in the US followed by India [6]. While the world fought the deadliest virus of the century with all the armamentarium available, the lack of optimal therapeutic strategies to combat the virus was acknowledged by the medical sector.

Along with repurposing of existing antivirals for COVID-19 management novel therapeutic solutions were explored at every phase of the pandemic, to eliminate the virus [7]. Till date, a number of non-specific broad-spectrum antivirals, specific antivirals, monoclonal antibodies, supportive therapies and vaccines have been developed [8]. Immunization campaigns were conducted globally to prevent spread of the disease. Many adverse events following administration of these novel vaccines including limb pain, fatigue, allergies, seizures and diarrhea were reported [9]. Studies also have shown that more than 70% of COVID survivors have mild to severe post COVID complications [10]. Conjointly, the adverse events of COVID and long COVID have severely reduced the health quotient of the population. Alternative complementary approaches can be of importance to gather evidence regarding early recovery from COVID19, halt the viral entry and manage chronic systems associated with Long COVID [11,12]. Herbal drugs are a less explored solution for combating the virus and its symptoms. Some of the commonly known phytochemicals present in herbal formulations including flavonoids, carotenoids, phenols and saponins, are known to have antimicrobial properties [13]. Thus, herbs offer promising alternatives in the quest for antimicrobial and antiviral treatments.

This study investigates a novel polyherbal formulation, NAOQ19, comprising 13 herbal elements against SARS CoV-2 virus. The formulation has been described in Supplementary Table 1. The formulation consists of well-known herbs like *Ashwangandha*, *Guduchi*, *Yashtimadhu*, *Haridra* with known antiviral, immunomodulatory and anti-allergic/anti-inflammatory activities [14]. Molecular docking studies on the above-mentioned herbs, have revealed seven potential inhibitors with binding energy of SARS-CoV-2 Viral N protein [15]. Another molecular docking study demonstrated the active phyto-constituents of *Bilwa* (*Aegle marmelos*) have stronger binding affinity against SARS-CoV-2 viral protein and better stability compared to Remdesivir [16]. *Vasaka* (*Adhatoda vasica*), another component of NAOQ19, known for its beneficial effects on respiratory disorders; has demonstrated multimodal therapeutic effects on the upper respiratory symptoms produced by COVID-19 [17]. Also documented are Vasaka’s antitussive, anti-inflammatory, antiviral and antioxidant pharmacological properties [18]. *Kalmegha* (*Andrographis paniculata*), has been studied for its antiviral properties against viruses such as chikungunya, dengue and influenza. It also possesses antithrombotic properties and helps in prevention of blood clot formation, which was one of the major concerns during COVID-19 [19].

During the height of the COVID pandemic, many classical antivirals with broad-spectrum antiviral activity often proved ineffective against newly evolved viral strains [20]. Thus, novel formulations with active components potentially complementing each other in increasing immunity and antiviral efficacy was the need of the hour. NAOQ19, a polyherbal formulation, was evaluated for its efficacy against COVID-19 using Vero E6 kidney cell lines [21]. However, there remains a notable gap in the literature regarding its interactions with human cell lines and the assessment of its effects in animal models. Therefore, the current study evaluates a comprehensive testing of polyherbal formulation, NAOQ19 starting from *invitro* evaluation in human cell lines for its specificity, to testing its efficacy in an animal model and finally in the clinical setting to find direct treatment benefits.

## 2. Materials and Methods

### 2.1 Intervention

This study investigates a novel polyherbal formulation, NAOQ19, comprising 13 herbal elements against SARS CoV-2 virus. The formulation has been described in Supplementary Table 1. The formulation consists of well-known herbs like *Ashwangandha*, *Guduchi*, *Yashtimadhu*, *Haridra* with known antiviral, immunomodulatory and anti-allergic/anti-inflammatory activities [14].

### 2.2 *In vitro* EC50 and CC50 determination

Three cell lines A549-ACE2, Vero E6, Huh 7.5.1 ACE2 TMPRSS2 were plated in 96 well plates at the density of 10,000 per well. After 24 hours, cells were infected with icSARS-CoV-2-nLuc (Bei NR54003) with an MOI of 0.1 in BSL-3. Subsequently the virus was washed off after 2 hours and drugs were added and incubated for 48 hours. Appropriate controls without drugs and DMSO-only groups were incorporated in the 96 well plates. After 48 hours, antiviral activity was determined by collecting the supernatant using nano glow luminescence method, where 100 µL nanoglow substrate (Nano-Glo Luciferase assay kit, Cat#N1130, Promega) was added in 100 µL of supernatant in a white plate. A promega microplate reader was under luminescence reading for EC50 determination. For determination of CC50, alamar blue viability assay was performed by adding 100 µL of warm medium in the above infected cell plates. This was followed by addition of 20 µL of Alamar Blue (AlamarBlue cell viability reagent, Cat#DAL1100, Invitrogen). Plates were then incubated for 2 hours at 37°C followed by plate reading (GM3000 Promega plate reader).

### 2.3 Efficacy in Syrian Golden Hamster *(In vivo* Assessment)

19 female hamsters were divided into 4 groups: viz. Mock control (uninfected), Infected Placebo control (untreated), infected positive control (Remdesivir), infected test (NAOQ19 treated). Animals (Female Syrian Golden Hamsters) were housed in the Foundation for Neglected Disease Research (FNDR) animal facilities. All the work was performed in compliance with the Control and Supervision of Experiments on Animals (CPCSEA), India regulations. The study was approved by FNDR’s Institutional Animal Ethics Committee (IAEC) with registration Number 2082/PO/Rc/S/19/CPCSEA. All the animals were kept in well ventilated cages and room conditions were well-maintained for their optimum living including food and water.

A viral concentration of 1× 10^5^ PFU SARS-CoV-2 (USAWA1/2020 viral isolate from ATCC) was administered intranasally into the hamsters. 15 mg/kg Remdesivir was administered intraperitoneally into the positive control group and 1000 mg/kg NAOQ19 was administered with an oral gavage into the study group daily in 24-hour intervals for 3 days. Body weights of hamsters were measured before and after the 3-day intervention. At initiation of the study the age of the hamster is 6-8 weeks and weighs 80-100 grams. At the end of the intervention, the animals were euthanized and sacrificed. The hamster lungs were then examined for any gross morphological and pathological changes.

To determine viral load, protocol by Abdelnabi et al. was followed [22]. Briefly, the lungs were homogenized and centrifuged to remove cellular debris. The supernatant was then serially diluted and plated onto a monolayer of Vero E6 cells. After an hour of incubation at 37°C, the cells were washed and incubated with DMEM: 2% Carboxy Methyl Cellulose Sodium Salt (1:1 ratio) for three days. The cells were then fixed with 4% formaldehyde for 30 minutes and after washing with 2x PBS at room temperature, the fixed cells were stained with 0.05% w/v crystal violet. Plaques were then counted at a dilution that gave readable counts and the PFU/lung was then determined by correcting the dilution factor.

### 2.4 Efficacy in human trials

A randomized double blinded clinical trial was conducted with 100 mild-moderate COVID-19 patients that tested positive on RT-PCR test (recruited 20 January 2022 - 23 February 2022)^1*^. Informed consent was obtained from all individuals included in this study. The study was approved by the ethical committee of the institute.

The eligibility criteria for the patients included an age range of 18-65 years with positive mild-moderate COVID-19 infection as reported on the RT-PCR test and ICMR guidelines. Patients with severe COVID-19 infection or those admitted in the Intensive Care Unit (ICU) were excluded from the study. Additionally, participants enrolled in other clinical trials or those unwilling to take the intervention were excluded from the study. Patients were randomized into a placebo and treatment group using a stratified randomization technique. NAOQ19 was administered at a dose of 1g (2 tablets) given three times a day after food. This was matched by 100% starch, placebo tablets, given to the placebo-control group. In addition to the intervention, patients were given standard of care treatment as described by Indian Council for Medical Research (ICMR) during the third wave of COVID in India [23]. This included antipyretics, antitussives and hydration medication along with azithromycin (500 mg, One Tablet, OD 5 days), vitamin C (one tablet, BD 7 days), zinc (one tablet, OD 7 days) as needed for mild patients. For moderate patients an injection of methylprednisolone (0.5 to 1 mg/kg) in 2 divided doses or an equivalent dose of dexamethasone for a duration of 5-10 days was administered as needed. This was supplemented with low-molecular weight heparin (0.6ml OD 10 days) if they were low on oxygen saturation levels.

Viral load was then measured by RT-PCR on days 3, 5 and 7 or until patients turned RT-PCR negative. Along with viral load, the inflammatory marker C-Reactive Protein was measured. Kidney Function Tests and Liver Function Tests were also carried out at entry and exit to evaluate safety of the drug. Further symptoms related to COVID were also monitored for assessing clinical recovery as mentioned by Singh et al [24] (refer Table 1A and Table 1B).

**Table1A:** Comparison of Kidney Function Test Parameters Between the Two Arms to evaluate the safety of NAOQ19 in clinical study.

| Time Points | Parameter | NAOQ19<br>Mean (SD) | Placebo<br>Mean (SD) |
| --- | --- | --- | --- |
| Day 1 | Urea | 24.26 (5.86) | 25.30 (12.45) |
| Day 7/exit |  | 24.92 (6.98) | 25.36 (6.4) |
| Day 1 | Creatinine | 0.90 (0.14) | 0.86 (0.12) |
| Day 7/exit |  | 0.89 (0.12) | 0.90 (0.12) |
| Day 1 | Uric Acid | 4.66 (0.98) | 4.96 (0.96) |
| Day 7/exit |  | 4.85 (0.87) | 5.12 (0.83) |

**Table1B:** Comparison of Liver Function Test Parameters Between the Two Arms to evaluate the safety of NAOQ19 in clinical study.

| Time Points | Parameter | NAOQ19<br>Mean (SD) | Placebo<br>Mean (SD) |
| --- | --- | --- | --- |
| Day 1 | S.G.O.T.(A.S.T.) | 33.91 (15.03) | 29.79 (9.45) |
| Day 7/exit |  | 30.52 (10.56) | 28.04 (8.98) |
| Day 1 | S.G.P.T. (A.L.T.) | 26.65 (9.93) | 20.63 (6.73) |
| Day 7/exit |  | 22.35 (7.33) | 23.17 (9.81) |
| Day 1 | Total Bilirubin | 0.91 (0.40) | 0.89 (0.39) |
| Day 7/exit |  | 0.71 (0.43) | 0.78 (0.39) |

**Primary outcomes** included reduction in viral load as assessed by RT-PCR on days 3, 5, and 7 (or until RT-PCR negativity), and clinical recovery from COVID-19 symptoms, evaluated using criteria described by Singh et al.

**Secondary outcomes** included changes in inflammatory marker C-Reactive Protein (CRP), and safety assessments based on kidney and liver function tests at baseline and study exit.

### 2.5 Statistical Methodology

#### i) Efficacy in Syrian Golden Hamster *(In vivo* Assessment)

Viral loads in the lungs were compared between the test and control groups by one-way analysis of variance (ANOVA) followed by Dennett’s multiple comparison using GraphPad Prism software (Version 9).

#### ii) Efficacy in human trials

Data entry was done in Microsoft Excel and analyzed using IBM SPSS Statistics, Version 23.0. Demographic variables were tabulated and represented using proportions (%). The analysis was performed using an intention to treat approach where all patients who were randomized were included in the analysis. The study sample was put through the Kolmogorov Smirnov test to check data normality. Chi-square test was used for comparisons between groups for categorical data. Quantitative data was presented with mean and SD and analyzed using Student’s T (paired and unpaired) test wherever applicable. Results from Kaplan Meier Analysis provided time to become asymptomatic between the two arms. The mean time amongst the two arms was tested for significance using the Log rank test. Significance was accepted with a two-sided p-value < 0.05 for the outcome measures.

## 3. Results

### 3.1 *In vitro* EC50 in Cell lines

To evaluate the potential effect of NAOQ19 on viral activity in vitro, an antiviral assay using the SARS-CoV-2-Nluc reporter virus was performed. Cells (Vero E6, Huh 7.5.1 ACE2 TMPRSS2 and A549-ACE2 TMPRSS2) were infected with the virus and treated with. Either NAOQ19 or the Remdesivir analog GS441524. Two days later, a cytotoxicity assay was performed with MOI 0.1 estimates the EC50 values. NAOQ19 inhibited SARS-CoV-2 viral load with EC50 values of 1.3 mg/ml (Vero E6), 0.8 mg/ml (A549-hACE2), and 1.7 mg/ml (Huh7.5.1 ACE2 TMPRSS2) without any apparent effect on cellular viability (Fig 1). GS441524 also inhibited viral load with EC50 of 11 µM in Vero E6, 11.4 µM in A549-hACE2 and 7.1 µM in Huh7.5.1 ACE2 TMPRSS2 cell lines, respectively without any significant effect on cell viability; these data are similar to those published earlier [25]. These results suggest that NAOQ19 has similar SARS-CoV-2 antiviral activity to GS441524 in vitro.

**Fig 1.**
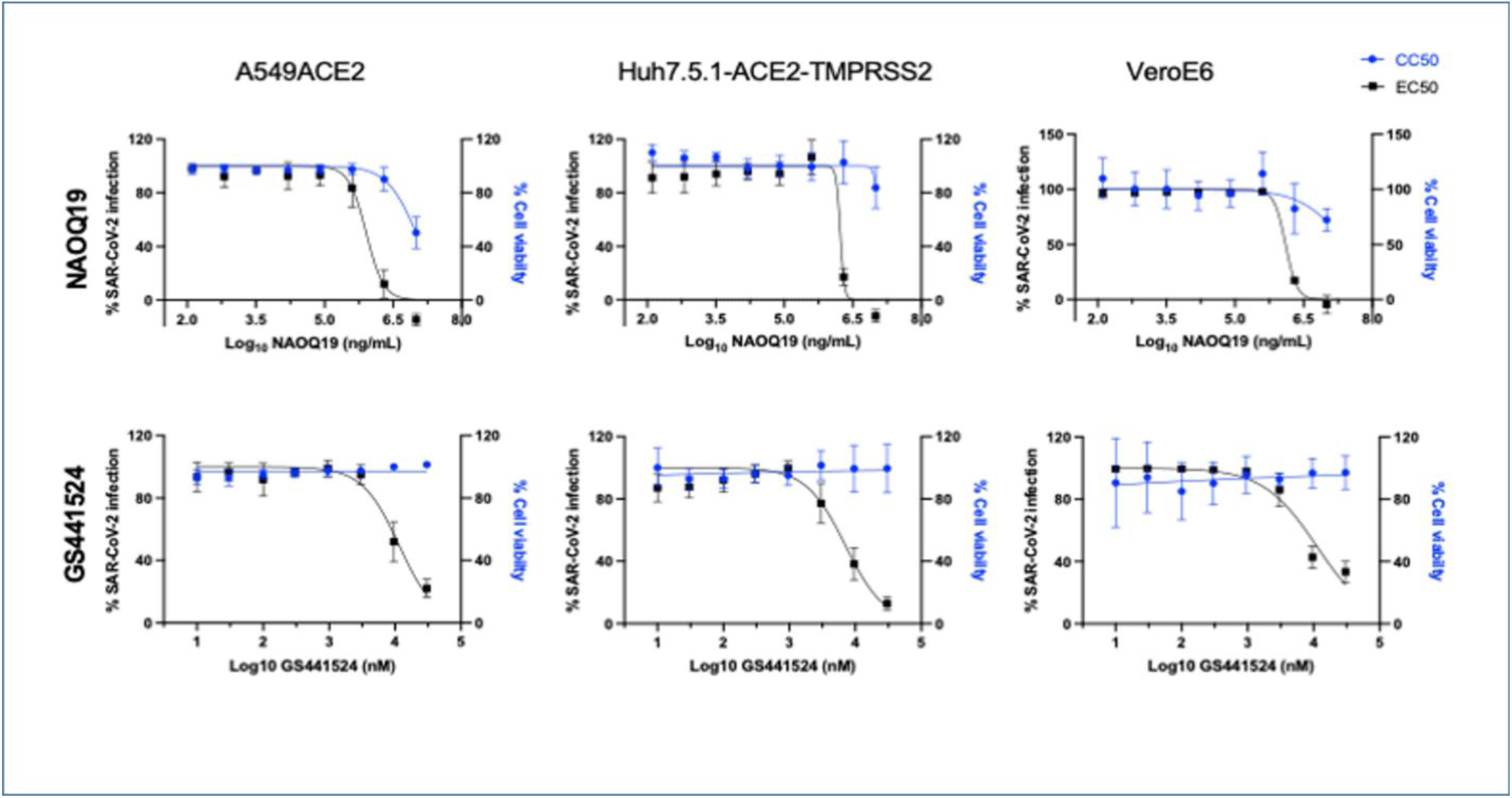

### 3.2 Efficacy in hamster model

A schematic representation of the sequence of events in the animal study with the results are presented in Fig 2.

**Fig 2.**
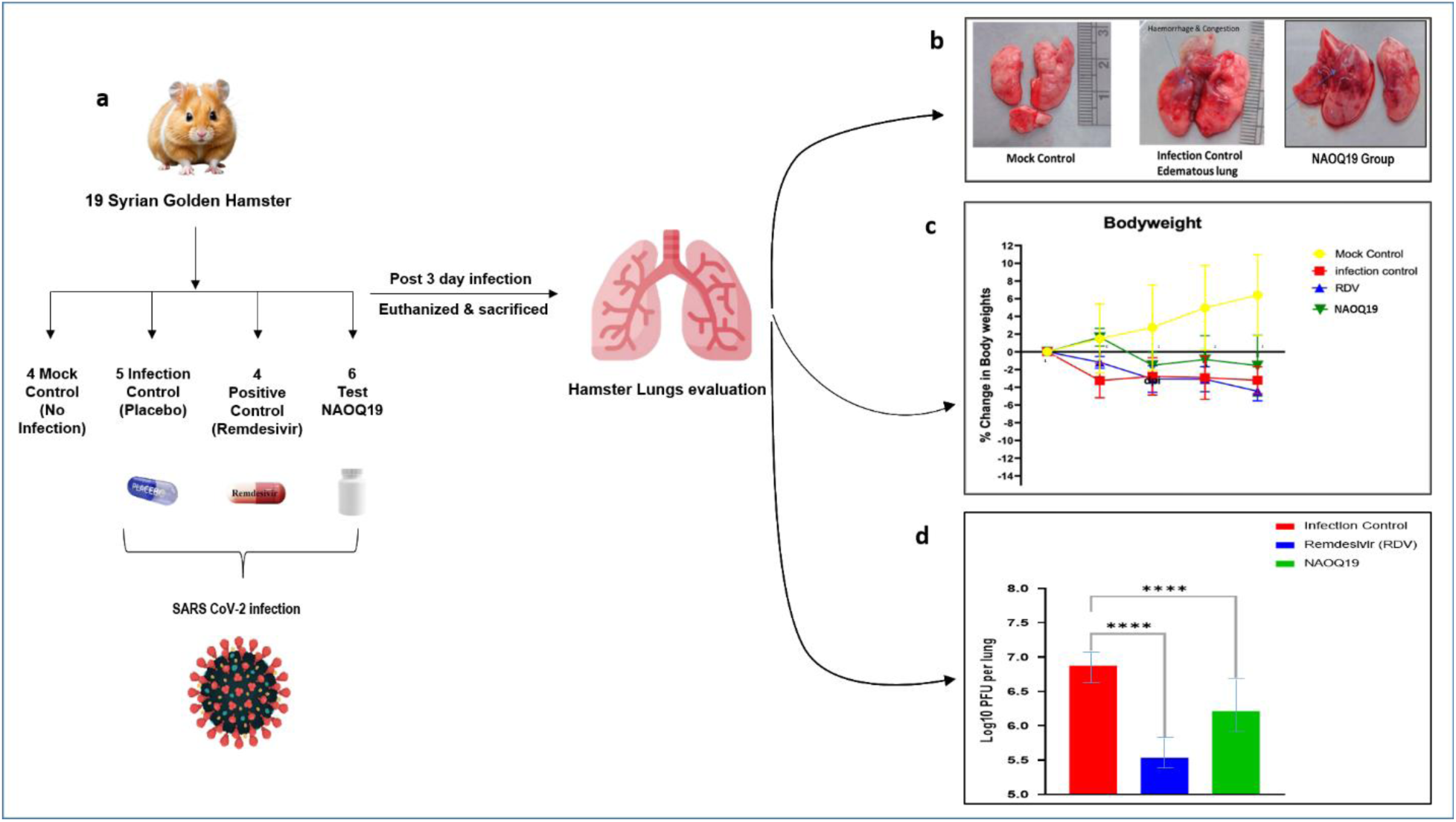

#### 3.2.1 Gross pathological examination

Gross pathological observations demonstrated normal hamster lungs in the non-infected group. The infected untreated group demonstrated severe edema and inflammation in all lobes (Fig 2B). The infected untreated group also had diffuse multi-focal hemorrhage and congestion. The NAOQ19 group showed improvement in lung edema, hemorrhage and congestion as compared to the untreated group. (Fig 2B).

The hamsters in uninfected mock control demonstrated an increase in their body weights during the study period. The infected but untreated hamsters showed a reduction in the body weight. NAOQ19 treated hamsters showed less body weight reduction when compared to the infection control and positive control. (Fig 2C).

#### 3.2.2 Viral load estimation

The test item NAOQ19 showed significant antiviral activity at 1000 mg/kg BID dosing in comparison to the untreated control group (Fig. 2D). Compared to the infected untreated group, the reduction of viral load in the NAOQ19 group was 78.2%. (Supplementary table 2).

### 3.3. Efficacy in human trials

Figure 3 represents the consort flow diagram of the clinical study population. Fig 4 A & B represents the demographic parameters among the study population. Both the arms were comparable with respect to age, gender and other vitals at baseline. No significant difference was noted between the two study groups.

**Fig 3.**
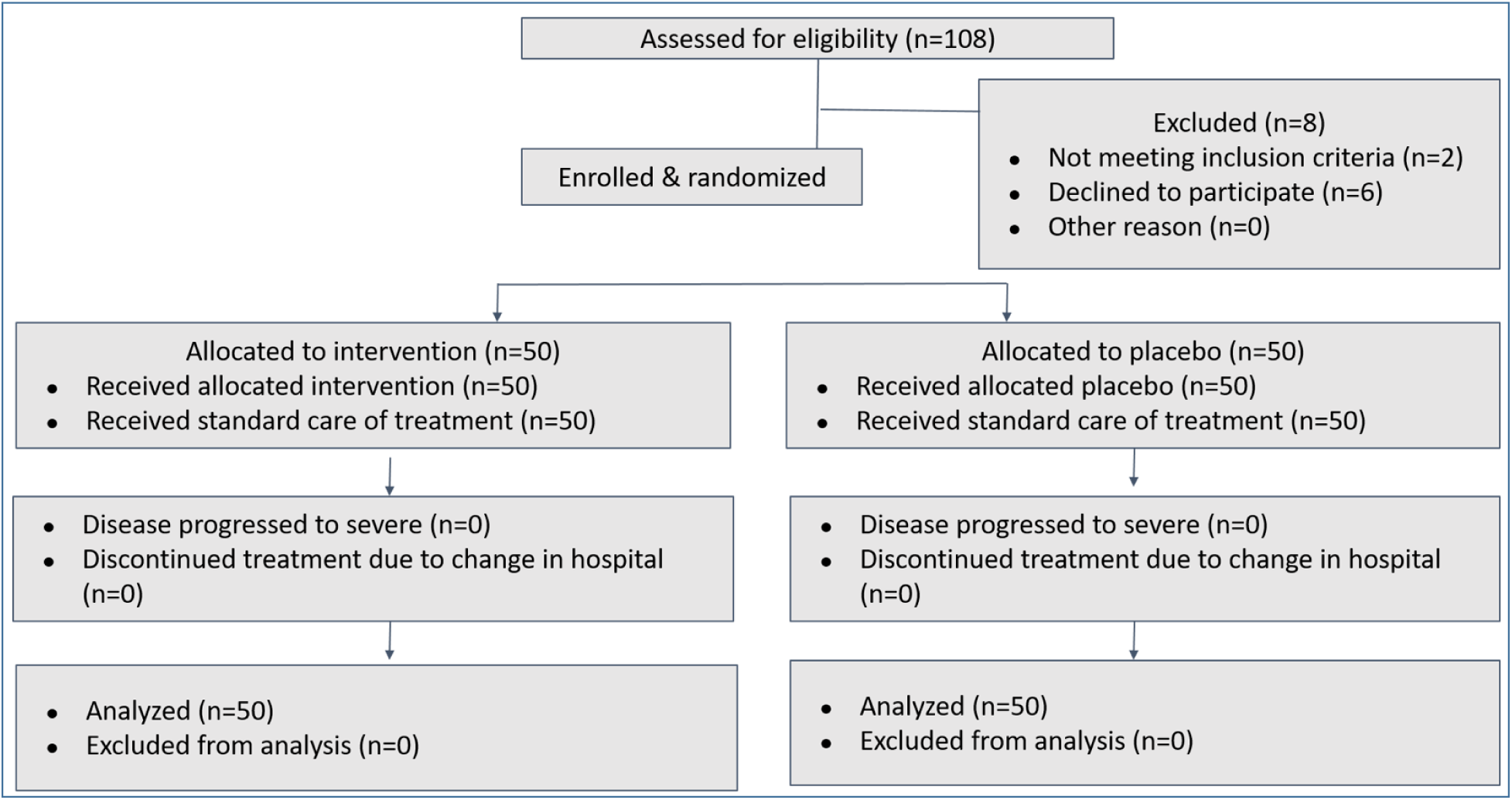

**Fig 4.**
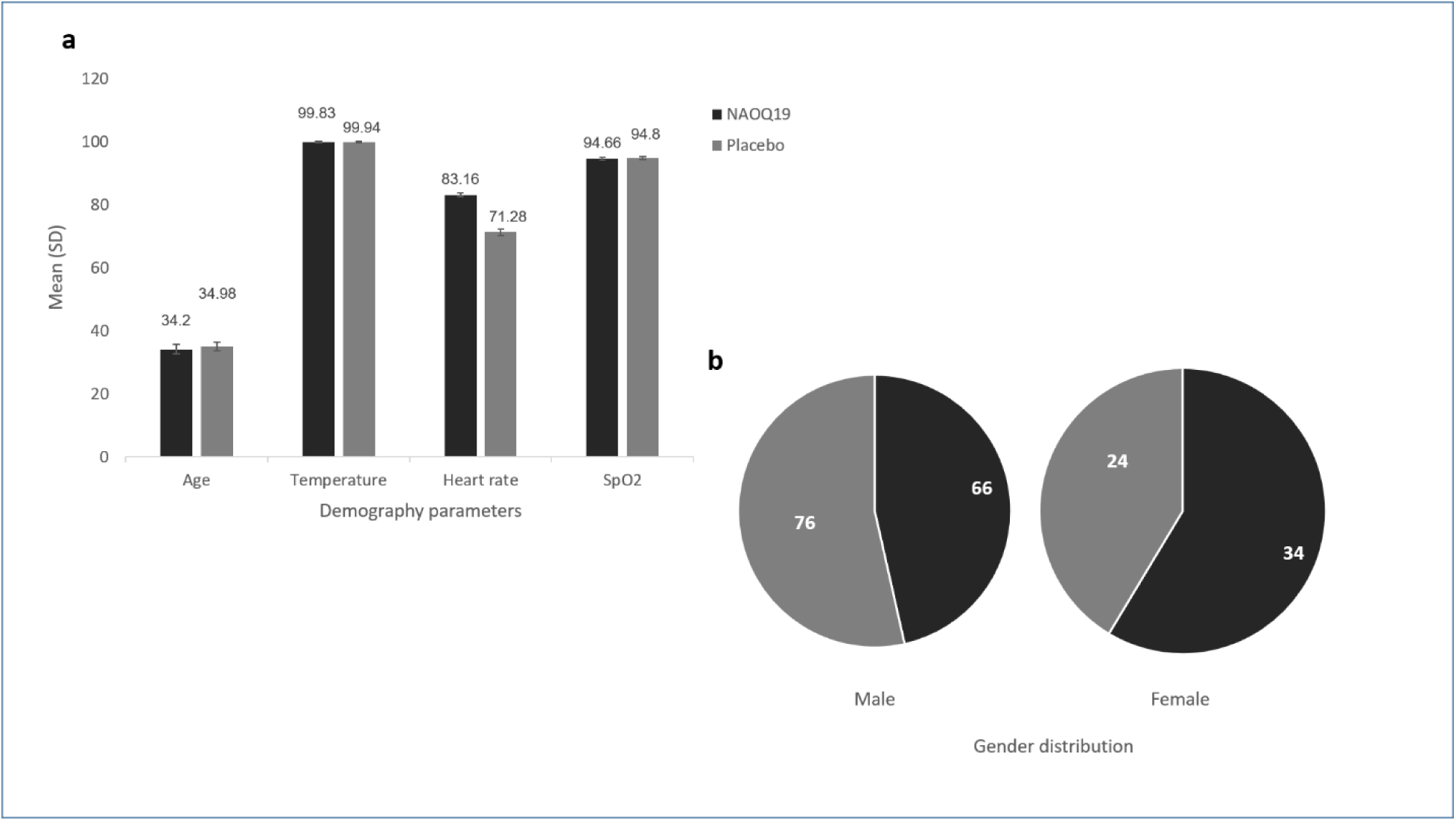

100 Mild-moderate COVID-19 patients were equally randomized into NAOQ19 group and placebo group. Both the group received the intervention along with standard of care till 7 days or until the patients turned RT-PCR negative as represented in Fig 5A. Fig 5B demonstrates the rate of recovery from COVID-19 in both the study arms as measured by RT-PCR test. Proportion of patients who were RT-PCR negative at day 3 were 34% in the NAOQ19 arm compared to none in the placebo arm. All the patients in the NAOQ19 arm turned RT-PCR negative by day 5 while only 34% patients turned negative in the placebo arm which was statistically significant.

**Fig 5.**
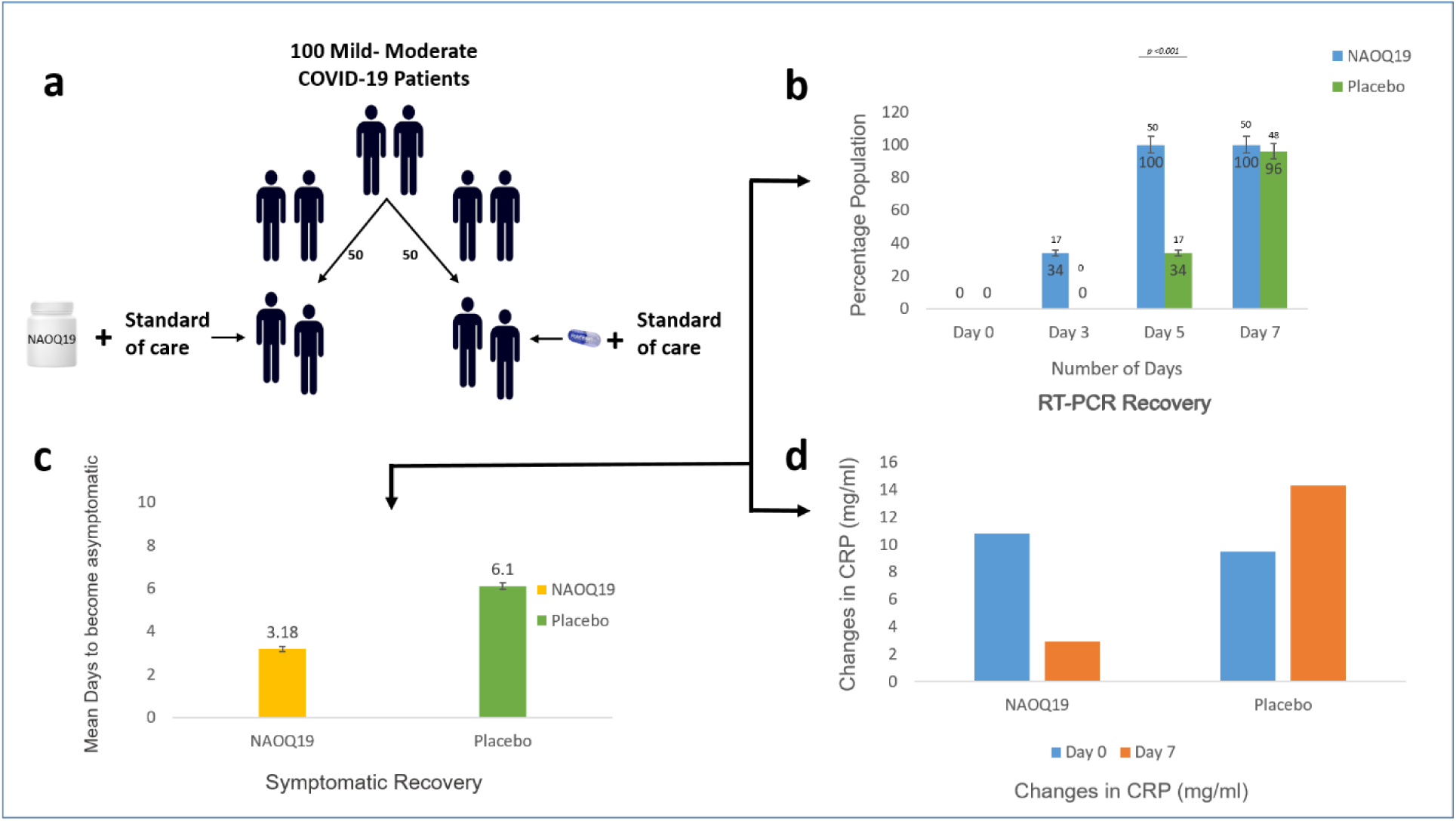

Fig 5C demonstrates the mean time to become asymptomatic between the two arms calculated using the Kaplan Meier analysis. The patients in the NAOQ19 arm recovered from symptoms in 3.18 days compared to 6.1 days in the placebo arm. This difference was found to be significant, p=<0.001 by Log Rank (Mantel-Cox) test. Fig 5D demonstrates the inflammatory marker C-reactive protein (CRP) values between the two arms during entry and exit. While no significant difference was seen in CRP at day 1 (p value=0.269) between the two study arms, median (Interquartile range) of CRP at day 7/exit in NAOQ19 was 7.65 (9.16) and in Placebo was 18.09 (19.69), respectively (p = 0.025). The CRP values had significantly reduced in the NAOQ19 group but increased in the placebo group at exit. No significant changes were observed in liver or kidney parameters from baseline to end line, indicating drug safety (Table 1A and 1B).

## 4. Discussion

This study comprehensively tests a new polyherbal Ayurvedic formulation, NAOQ19 for its efficacy against SARS-CoV-2. NAOQ19 was found to be effective in eliminating the virus *in vitro*, *in vivo* (hamster animal model) and in COVID-19 patients. Pre-clinical tests of NAOQ19 using Syrian Golden Hamsters showed that the oral supplementation of the drug is effective in reducing viral load and symptoms. Lower body weight reduction of the COVID-19-infected hamsters in the NAOQ19 group also showed that host health recovery could be faster. In the final phase, NAOQ19 was clinically tested on hospitalized COVID-19 patients. It was found that within 5 days of using NAOQ19 in conjunction with standard of care; 100% of patients tested RT-PCR negative for COVID-19, thereby significantly shortening the duration of the infection. At the time, the main cause of many COVID-19 hospital deaths was due to increased inflammation. Notably, the NAOQ19 group exhibited significantly reduced levels of CRP, a biomarker for inflammation, demonstrating its anti-inflammatory effects. The results of our study is congruous to the previous clinical study conducted that were NAOQ19 group demonstrated early recovery and higher proportion of participants [26–28]. To our knowledge, this is the first study to comprehensively test a polyherbal Ayurvedic formulation, from preclinical to translational stages, for its efficacy against COVID-19.

For decades, traditional medicine has been explored in pharmacology to treat long-standing diseases, continually reaffirming their importance in the present era. Due to the shortage of effective drugs caused by increased antimicrobial resistance and lengthy drug manufacturing process, the pharmaceutical industry is increasingly turning towards better natural and herbal remedies. The present study evaluates a polyherbal formulation including well-known antimicrobials such as *Ashwagandha*, *Guduchi* and *Tulasi* [26]. The results of the present study demonstrated no side effects alongside an improved recovery rate. Amidst the high morbidity rates during COVID-19, the pharmaceutical industry thrived by repurposing existing antivirals [27]. However, several studies demonstrated invasive fungal infections and superinfections among the moderate to severe infected COVID-19 patients after the use of corticosteroids [28]. Also, hyperglycemia, hypertension, fluid retention and cataract were some of the side effects with increased doses of dexamethasone corticosteroids [29]. Remdesivir, a popular repurposed antiviral, proved to be effective against COVID-19 but has also reported several side effects in clinical trials. Most common side effects included gastrointestinal upset and elevated levels of liver enzymes indicating an inflammation or damage to the liver cells [30]. The other two antimalarial drugs used in treatment of COVID-19, Hydroxychloroquine and Chloroquine, have also demonstrated several side effects in individuals such as cardiomyopathy, muscle weakness, seizures, myasthenia gravis and neuropathy under long term usage [31]. It was interesting to note that our clinical trial results not only demonstrated significant viral elimination but also helped alleviate the clinical symptoms caused by the disease. Moreover, the study reported no adverse events or side effects in either the animal model or human trial validating its safety and efficacy.

Another growing issue in the pharmaceutical industry is that of antimicrobial resistance (AMR). Currently AMR is one of the leading global concerns causing more than 4.75 million deaths globally [32]. As we observe a decrease in the effectiveness of antimicrobials drugs due to their overuse and inappropriate application leading to antimicrobial resistance, complementary and alternative medicine, including polyherbal formulations, can provide a significant advantage. Although there was no antimicrobial resistance reported for the novel COVID-19 infection, secondary nosocomial infections and superinfections developed as a result of AMR. A rapid increase in multidrug-resistant organisms was noted due to the high rate of antimicrobial usage and drug repurposing [33]. In the fight against COVID-19, herbal medicines have proven to be a valuable antimicrobial alternative, effectively combating the issue of antimicrobial resistance [34]. Indian, Chinese and several other governments issued national guidelines recommending the use of polyherbal formulations alongside standard therapy to combat the pandemic [35,36].

Compared to several other single herbal extracts and standardized concoctions used in the region, NAOQ19 can offer an added advantage. The individual herbal extracts used in NAOQ19 have a well-researched background as anti-viral and anti-inflammatory agent [37]. However, the compounded effect of the herbs in the formulation is yet to be studied to understand the mechanistic action of NAOQ19. Although the pandemic is gradually subsiding, its shadow lingers in the form of post-COVID complications. As quoted in the clinical study, its effectiveness in addressing the various symptomatic challenges posed by the virus is well established [38]. The results of the animal model study demonstrate the formulation’s benefit in reducing weight loss incidence post-viral attack, while the clinical trial results illustrate a reduction in inflammatory markers in the NAOQ19 group, a benefit absent in the placebo group. Studies cite high inflammation post COVID as one of the major reasons for post COVID symptoms including cardiomyopathy [39,40]. Future work on NAOQ19 would include its efficacy in abating post COVID complications among the previously infected individuals. Additionally, this formulation has only been studied for its efficacy against the SARS-CoV-2 virus. Testing the polyherbal formulation against other viruses like Influenza, a significant public health concern in many parts of the world including the United States [41], could provide an added benefit to the medical industry.

## 5. Conclusion

A large number of herbal supplements have been tried and tested against COVID-19. While some improved the general health of patients, they were ineffective for aged cohorts or immunocompromised groups, who eventually succumbed to the pandemic. This study tested a new polyherbal Ayurvedic formulation, NAOQ19, through a) *in vitro* analysis in three relevant cell lines, b) pre-clinical studies with a mammalian model, and c) clinical trials with COVID-19 patients. NAOQ19 demonstrated specific antiviral activity against SARS-CoV-2 in all three models. Additionally, NAOQ19 reduced inflammation, accelerated the conversion to RT-PCR negative, and consequently shortened the duration of hospital stays. Integrative health approaches can offer more personalized and engaging health interventions which can combat public health fatigue.

## Data Availability

All data produced in the present study are available upon reasonable request to the authors

## Footnotes

1* (Further details in Supplementary file)

## References

1. Piret J, Boivin G. Pandemics Throughout History. Front. Microbiol. 2021;11:631736.

2. Edwards AM, Baric RS, Saphire EO, Ulmer JB. Stopping pandemics before they start: Lessons learned from SARS-CoV-2. Science 2022;375:1133–9.

3. Coronavirus disease (COVID-19) pandemic [Internet]. [cited 2025 Jul 30];Available from: https://www.who.int/europe/emergencies/situations/covid-19

4. Hiscott J, Alexandridi M, Muscolini M, Tassone E, Palermo E, Soultsioti M, et al. The global impact of the coronavirus pandemic. Cytokine & Growth Factor Reviews 2020;53:1–9.

5. Liu W, Huang Z, Xiao J, Wu Y, Xia N, Yuan Q. Evolution of the SARS-CoV-2 Omicron Variants: Genetic Impact on Viral Fitness. Viruses 2024;16:184.

6. Paul S. Analyzing the attitude of Indian citizens during the second wave of COVID-19: A text analytics study. International Journal of Disaster Risk Reduction 2022;79:103161.

7. Raghav PK, Mann Z, Ahluwalia SK, Rajalingam R. Potential treatments of COVID-19: Drug repurposing and therapeutic interventions. Journal of Pharmacological Sciences 2023;152:1–21.

8. Murakami N, Hayden R, Hills T, Al-Samkari H, Casey J, Del Sorbo L, et al. Therapeutic advances in COVID-19. Nat Rev Nephrol 2023;19:38–52.

9. Kouhpayeh H, Ansari H. Adverse events following COVID-19 vaccination: A systematic review and meta-analysis. International Immunopharmacology 2022;109:108906.

10. Koc HC, Xiao J, Liu W, Li Y, Chen G. Long COVID and its Management. Int. J. Biol. Sci. 2022;18:4768–80.

11. Paudyal V, Sun S, Hussain R, Abutaleb MH, Hedima EW. Complementary and alternative medicines use in COVID-19: A global perspective on practice, policy and research. Research in Social and Administrative Pharmacy 2022;18:2524–8.

12. Wanjarkhedkar P, Sarade G, Purandare B, Kelkar D. The Post-COVID 19 long term surveillance study sequel to an add-on Ayurveda regimen. Journal of Ayurveda and Integrative Medicine 2022;13:100575.

13. Zrig A. The Effect of Phytocompounds of Medicinal Plants on Coronavirus (2019-NCOV) Infection. Pharm Chem J 2022;55:1080–4.

14. Singh R, Goel S, Bourgeade P, Aleya L, Tewari D. Ayurveda Rasayana as antivirals and immunomodulators: potential applications in COVID-19. Environ Sci Pollut Res 2021;28:55925–51.

15. Karakkadparambil Sankaran S, Nair AS. Molecular dynamics and docking studies on potentially active natural phytochemicals for targeting SARS-CoV-2 main protease. Journal of Biomolecular Structure and Dynamics 2023;41:6459–75.

16. Joshi C, Chaudhari A, Joshi C, Joshi M, Bagatharia S. Repurposing of the herbal formulations: molecular docking and molecular dynamics simulation studies to validate the efficacy of phytocompounds against SARS-CoV-2 proteins. Journal of Biomolecular Structure and Dynamics 2022;40:8405–19.

17. Gheware A, Dholakia D, Kannan S, Panda L, Rani R, Pattnaik BR, et al. Adhatoda Vasica attenuates inflammatory and hypoxic responses in preclinical mouse models: potential for repurposing in COVID-19-like conditions. Respir Res 2021;22:99.

18. Shoaib A. A systematic ethnobotanical review of Adhatoda vasica (L.), Nees. Cell Mol Biol (Noisy-le-grand) 2022;67:248–63.

19. Intharuksa A, Arunotayanun W, Yooin W, Sirisa-ard P. A Comprehensive Review of Andrographis paniculata (Burm. f.) Nees and Its Constituents as Potential Lead Compounds for COVID-19 Drug Discovery. Molecules 2022;27:4479.

20. Bolinger AA, Li J, Xie X, Li H, Zhou J. Lessons learnt from broad-spectrum coronavirus antiviral drug discovery. Expert Opinion on Drug Discovery 2024;19:1023–41.

21. Kanchibhotla D, Subramanian S, Ravi Kumar RM, Venkatesh Hari KR, Pathania M. An In-vitro evaluation of a polyherbal formulation, against SARS-Cov-2. Journal of Ayurveda and Integrative Medicine 2022;13:100581.

22. Abdelnabi R, Foo CS, Jochmans D, Vangeel L, De Jonghe S, Augustijns P, et al. The oral protease inhibitor (PF-07321332) protects Syrian hamsters against infection with SARS-CoV-2 variants of concern. Nat Commun 2022;13:719.

23. RevisedNationalClinicalManagementGuidelineforCOVID1931032020.pdf.

24. Singh H, Srivastava S, Yadav B, Rai AK, Jameela S, Muralidharan S, et al. AYUSH-64 as an adjunct to standard care in mild to moderate COVID-19: An open-label randomized controlled trial in Chandigarh, India. Complementary Therapies in Medicine 2022;66:102814.

25. Amirian ES, Levy JK. Current knowledge about the antivirals remdesivir (GS-5734) and GS-441524 as therapeutic options for coronaviruses. One Health 2020;9:100128.

26. Alam S, Sarker MdMR, Afrin S, Richi FT, Zhao C, Zhou JR, et al. Traditional Herbal Medicines, Bioactive Metabolites, and Plant Products Against COVID-19: Update on Clinical Trials and Mechanism of Actions. Front. Pharmacol. 2021;12:671498.

27. Ng YL, Salim CK, Chu JJH. Drug repurposing for COVID-19: Approaches, challenges and promising candidates. Pharmacology & Therapeutics 2021;228:107930.

28. Søvik S, Barratt-Due A, Kåsine T, Olasveengen T, Strand MW, Tveita AA, et al. Corticosteroids and superinfections in COVID-19 patients on invasive mechanical ventilation. Journal of Infection 2022;85:57–63.

29. Langarizadeh MA, Ranjbar Tavakoli M, Abiri A, Ghasempour A, Rezaei M, Ameri A. A review on function and side effects of systemic corticosteroids used in high-grade COVID-19 to prevent cytokine storms. EXCLI Journal; 20:Doc339; ISSN 1611-2156 [Internet] 2021 [cited 2025 Jul 30];Available from: https://www.excli.de/index.php/excli/article/view/3196

30. Aleem A, Mahadevaiah G, Shariff N, Kothadia JP. Hepatic manifestations of COVID-19 and effect of remdesivir on liver function in patients with COVID-19 illness. Baylor University Medical Center Proceedings 2021;34:473–7.

31. Carvalho AADS. Side Effects of Chloroquine and Hydroxychloroquine on Skeletal Muscle: a Narrative Review. Curr Pharmacol Rep 2020;6:364–72.

32. Antimicrobial resistance [Internet]. [cited 2025 Jul 30];Available from: https://www.who.int/news-room/fact-sheets/detail/antimicrobial-resistance

33. Sharma S, Barman P, Joshi S, Preet S, Saini A. Multidrug resistance crisis during COVID-19 pandemic: Role of anti-microbial peptides as next-generation therapeutics. Colloids and Surfaces B: Biointerfaces 2022;211:112303.

34. Van Wietmarschen HA. Integrative approaches to antimicrobial resistance. European Journal of Integrative Medicine 2020;39:101191.

35. Kotecha R. The journey with COVID-19: Initiatives by Ministry of AYUSH. Journal of Ayurveda and Integrative Medicine 2021;12:1–3.

36. Zhao Z, Li Y, Zhou L, Zhou X, Xie B, Zhang W, et al. Prevention and treatment of COVID-19 using Traditional Chinese Medicine: A review. Phytomedicine 2021;85:153308.

37. Bhardwaj P, Ganapathy K, Pathania M, Naveen KH, Charan J, Dutta S, et al. Effectiveness of ayurvedic formulation, NAOQ19 along with standard care in the treatment of mild-moderate COVID-19 patients: A double blind, randomized, placebo-controlled, multicentric trial. Journal of Ayurveda and Integrative Medicine 2023;14:100778.

38. Das SK, Dash DP, Panda PK, Sadana S, M Ravi Kumar R, Hari K V, et al. The Efficacy of a Plant Based Formulation in the Symptomatic Management of Mild COVID-19 Cases: A Double Blind, Randomized Controlled Trial. Arch Clin Med Case Rep [Internet] 2023 [cited 2025 Jul 30];07. Available from: https://www.fortunejournals.com/articles/the-efficacy-of-a-plant-based-formulation-in-the-symptomatic-management-of-mild-covid19-cases-a-double-blind-randomized-controlled.html

39. Arun A, Subramanian S, Kanchibhotla D. Efficacy of polyherbal formulation along with standard care of treatment in early recovery of COVID-19 patients: a randomized placebo-controlled trial. Beni-Suef Univ J Basic Appl Sci 2023;12:103.

40. Maamar M, Artime A, Pariente E, Fierro P, Ruiz Y, Gutiérrez S, et al. Post-COVID-19 syndrome, low-grade inflammation and inflammatory markers: a cross-sectional study. Current Medical Research and Opinion 2022;38:901–9.

41. Ali ST, Lau YC, Shan S, Ryu S, Du Z, Wang L, et al. Prediction of upcoming global infection burden of influenza seasons after relaxation of public health and social measures during the COVID-19 pandemic: a modelling study. The Lancet Global Health 2022;10:e1612–22.

