## Supplementary table 1, Supplementary table 2, Supplementary file for "Preclinical and Clinical Study of Efficacy of NAOQ19 against SARS-COV2: A Comprehensive Evaluation"

| S.No | Name of the herb | Scientific Name | Part Used | Nature of herb | Quantity (mg) |
|---|---|---|---|---|---|
| 1 | Ashwagandha | *Withania Somnifera* | Root | Fine Powder, Extract | 30 |
| 2 | Bilwa | *Aegle marmelos* | Leaf | Fine Powder | 30 |
| 3 | Yashtimadhu | *Glycyrrhiza glabra* | Root | Fine Powder, Extract | 20 |
| 4 | Rasna | *Pluchea lanceolata* | Leaf | Fine Powder | 30 |
| 5 | Vasaka | *Adhatoda vasica* | Leaf | Fine Powder, Extract | 25 |
| 6 | Pippali | *Piper longum* | Fruit | Fine Powder | 30 |
| 7 | Bhumiamla | *Phyllantus fraternus* | Plant | Fine Powder, Extract | 35 |
| 8 | Kalmegh | *Andrographis paniculata* | Whole plant | Fine Powder | 30 |
| 9 | Saptaparna | *Alstonia scholaris* | Stem bark | Fine Powder | 30 |
| 10 | Haridra | *Curcuma longa* | Rhizome | Fine Powder | 25 |
| 11 | Patha | *Cissampelous pareira* | Root | Fine Powder | 25 |
| 12 | Tulasi | *Ocimum sanctum* | Whole plant | Fine Powder | 20 |
| 13 | Guduchi | *Tinospora cordifolia* | Stem | Fine Powder | 20 |

**Supplementary table 1: Composition of NAOQ19.** The list of ingredients in NAOQ19 with the proportion of the drug has been provided.

| Group | No of plaques 1 | 2 | Avg | Dilution Factor | Vol of sample used for infection (mL) | PFU/mL | PFU/Lung | Log PFU/Lung | Avg PGU /Lung | Log reduction |
|---|---|---|---|---|---|---|---|---|---|---|
| Mock Infection | 0 | 0 | 0 | 10 | 0.05 | 0 | 0 | 0.00 | 0.00 | |
| | 0 | 0 | 0 | 10 | 0.05 | 0 | 0 | 0.00 | | |
| | 0 | 0 | 0 | 10 | 0.05 | 0 | 0 | 0.00 | | |
| | 0 | 0 | 0 | 10 | 0.05 | 0 | 0 | 0.00 | | |
| Infection Control | 19 | 18 | 18.5 | 10000 | 0.05 | 3700000 | 7400000 | 6.87 | 6.88 | |
| | 18 | 14 | 16 | 10000 | 0.05 | 3200000 | 6400000 | 6.81 | | |
| | 22 | 25 | 23.5 | 10000 | 0.05 | 4700000 | 9400000 | 6.97 | | |
| | 16 | 18 | 17 | 10000 | 0.05 | 3400000 | 6800000 | 6.83 | | |
| | 21 | 19 | 20 | 10000 | 0.05 | 4000000 | 8000000 | 6.90 | | |
| RDV | 5 | 8 | 6.5 | 1000 | 0.05 | 130000 | 260000 | 5.41 | 5.53 | 1.34 |
| | 12 | 10 | 11 | 1000 | 0.05 | 220000 | 440000 | 5.64 | | |
| | 9 | 11 | 10 | 1000 | 0.05 | 200000 | 400000 | 5.60 | | |
| | 6 | 9 | 7.5 | 1000 | 0.05 | 150000 | 300000 | 5.48 | | |
| NAOQ19 | 8 | 4 | 6 | 10000 | 0.05 | 1200000 | 2400000 | 6.38 | 6.22 | 0.66 |
| | 15 | 18 | 16.5 | 1000 | 0.05 | 330000 | 660000 | 5.82 | | |

| | | | | | | | |
|---|---|---|---|---|---|---|---|
| 5 | 4 | 4.5 | 10000 | 0.05 | 900000 | 1800000 | 6.26 |
| 17 | 19 | 18 | 1000 | 0.05 | 360000 | 720000 | 5.86 |
| 6 | 9 | 7.5 | 10000 | 0.05 | 1500000 | 3000000 | 6.48 |
| 10 | 6 | 8 | 10000 | 0.05 | 1600000 | 3200000 | 6.51 |

**Supplementary table 2:** Viral Load estimation using the plaque formation units (PFU) per lung in the Syrian Golden Hamsters.

### Supplementary Methods

#### Animal Study

*1.1 Concentration and Vehicle for NAOQ19 intervention in Syrian Golden Hamsters*

NAOQ19 intervention was given via oral gavage post 24 hours of infection. The intervention was given using saline at a dose of 1000mg/kg twice daily for 3 days.

*1.2 Concentration and Vehicle for Remdesivir intervention in Syrian Golden Hamsters*

Remdesivir intervention was given intraperitoneally post 24 hours of infection. The intervention was given using 5% DMSO/5%Ethanol/ 40%PEG 300/50% Normal saline at a dose of 15mg/kg daily for 3 days.

#### Clinical Trials

*2.1 Trial site*:

The participants were recruited from a 450 bedded tertiary care Covid care center (Govt. Institute for Medical Sciences, New Delhi) in Uttar Pradesh, the north of India. The site was well equipped with facilities such as RT-PCR testing and other laboratory based tests and plasma therapy.

The patients were allocated into the placebo or intervention group as per the randomization and inflow of the patients. Each patient received a bottle of 90 tablets. Both the NAOQ19 and placebo bottles were packaged in identical white containers to avoid any bias. The treatment compliance was monitored by recording the medicines used in the containers during each scheduled visit. In case the patient missed a dose, the subsequent dose was continued while the missed dose was reported in the Clinical Report Form (CRF).

*2.2 Treatment group: NAOQ19 drug:*

NAOQ19 is a polyherbal formulation with 13 herbal constituents well-known for its antiviral and anti-inflammatory properties. The formulation contains a blend of fine powders and aqueous extracts for some of its ingredients as mentioned in Supplementary Table 1. The formulation was prepared at Sriveda Sattva Private Ltd, Bangalore, a GMP certified organization. All the herbal extracts were subjected to quality control in its crude form. After passing the quality control, all the ingredients were blended with excipients, granulated, dried and compressed. The tablets were then packed using standard procedures. The formulation was approved by the Ministry of AYUSH, India for mild-moderate COVID-19 patients along with standard care of therapy. The dosage for the present study was two tablets (1mg), 3 times a day after food along with standard care of therapy.

*2.3 Control group*: Placebo:

Tablets that matched color, size and shape with that of NAOQ19 was used as placebo. The tablets were made of 100% starch and passed quality control test before packing into identical bottles. The dosage for the present study was two tablets, 3 times a day after food along with standard care of therapy.

*2.4 Randomization and Blinding*

The patients enrolling in the study were randomly assigned to the intervention group or the placebo group in the ratio 1:1. Stratified randomization was performed based on the age and onset of symptoms. The participants were informed of their respective intervention with an envelope that was sequentially numbered, sealed and distributed. To avoid bias, randomization was performed by intern staff who was not involved in the study. The outcome assessor, statistician was also blinded to the intervention.

*2.5 Sample size justification*

The sample size was calculated based on the study of Devapura G, et al. (1), where the treatment group witnessed 100% recovery by day 7, while it was 60% in the placebo group. Taking these values as reference, the minimum required sample size with 99% power of study and 1% level of significance was 37 patients in each study group. To reduce margin of error, a total sample size of 100 (50 patients per group) patients were enrolled.

**Reference**


1. Devpura G, Tomar BS, Nathiya D, Sharma A, Bhandari D, Haldar S, Balkrishna A, Varshney A. Randomized placebo-controlled pilot clinical trial on the efficacy of ayurvedic treatment regime on COVID-19 positive patients. Phytomedicine. 2021 Apr 1;84:153494.
